# Psychosocial and psychological Interventions’ effectiveness among Nurses in Intensive Care Units caring for pediatric patients: A Systematic Review and Meta-Analysis

**DOI:** 10.1101/2022.03.30.22273187

**Authors:** Mi-Hyang Choi, Mi-Soon Lee

## Abstract

**Purpose:** This review aimed to evaluate the effectiveness of psychosocial and psychological interventions in nurses among intensive care units caring for pediatric patients.

**Methods:** This study was applied to the Participants, Intervention, Comparisons, Outcomes, Timing of Outcome Measurement, Settings, Study Design and PubMed, EMBASE, CINAHL databases were searched. To estimate the effect size, a meta-analysis of the studies was performed using the RevMan 5.3 program. The effect size used was the standardized mean difference.

**Results:** Of 1,630 studies identified, 4 met the inclusion criteria, and 3 studies were used to estimate the effect size of psychosocial and psychological interventions. The primary outcome variable of these studies was stress. The effect of the intervention program on stress was also found to have no effect in individual studies, and the overall effect size was not statistically significant (SMD = -0.06; 95% CI: -0.33, 0.20; Z = 0.48, p = 0.630). However, according to the individual literature included in this study, after the stress management program was applied as a group, a significant stress reduction was shown in the experimental group (p = 0.021).

**Conclusions:** These results show that psychosocial and psychological interventions were effective in stress management by a group approach. Therefore, it is necessary to develop psychosocial support interventions for stress management of nurses among intensive care units caring for pediatric patients more diversely.

## 1. Introduction

Due to the COVID-19 pandemic, the psychological stress faced by hospital medical staff is higher than ever. The prevalence of stress among medical staff was 45%, which was higher than the prevalence of anxiety (25.8%) and depression (24.3%) [1]. Such stressors were relatively more common in nurses than in other healthcare workers [2, 3]. Nursing is a profession with higher psychosocial stress than other occupations as it involves direct care of patients. Among them, nurses working in intensive care units (ICU) showed higher psychosocial stress due to the severity of the patients’ diseases. Particularly, ICU nurses caring for pediatric patients felt rewarded at work but displayed high-stress levels. Further, 55.8% of ICU nurses caring for pediatric patients claimed to be stressed frequently [4]. Moreover, they showed middle-high or higher stress responses [5] and had a more significant burden than nurses providing care for terminal adult patients [6]. Nurses providing care for pediatric patients are faced with elevated psychological stress arising from various factors, including the burden from caring for a weak child, patients’ death, dealing with various reactions of the parents [7, 8], having to witness patients’ pain and suffering while spending time with them, irregular work shifts, inappropriate emergency management and insufficient experience, postmortem management for pediatric patients [9], ethical dilemmas encountered during work, staffs’ work incompetence [10], and inappropriate staffing [4].

Stress includes both distress and eustress [11, 12], and most stress experienced by ICU nurses caring for pediatric patients is high moral distress. The moral distress score among ICU nurses caring for pediatric patients in Canada was above 100. This is higher than the average of 79 and the moral distress scores of physicians. Particularly, nurses providing life-sustaining treatment for children with no survival hope due to a request from their family showed a higher level of moral distress [13]. Additionally, other factors caused moral stress, including communication with the patients’ parents and treatment team, relationship with the treatment team for patient-caring, continuous use of invasive treatment modalities for a dying or suffering child, the training process to become sufficiently skilled, and higher work-related challenges and sensitivity than other nursing units because of the responsibility of correctly manipulating several machines [14]. These results suggest that ICU nurses caring for pediatric patients demonstrate higher stress levels than other nursing units. Moreover, they work in an environment that readily triggers moral distress.

High-stress levels in ICU nurses caring for pediatric patients threaten their mental health. This demoralizes them, induces fatigue, and contributes to inappropriate care, leading to job dissatisfaction, burnout, and turnover [9, 15, 16]. Particularly, moral distress induces negative emotions such as grief, anxiety, helplessness, and dissatisfaction among nurses [8, 10, 13], leading to burnout and uncertainty among ICU nurses caring for pediatric patients [16]. Moreover, it contributes to the deterioration of the quality of nursing care for pediatric patients [8, 17]. Nurses experiencing such negative emotions tend to avoid facing them [18] because they believe that expressing their emotions is unprofessional [19]. Thus, the aggressive management and prevention of stressors developed from work are critical for ICU nurses caring for pediatric patients working in a highly sensitive environment. When appropriately implemented, this can enhance the quality of nursing care for pediatric patients.

Psychosocial and psychological interventions include psychoeducational strategies, cognitive behavioral therapy, interpersonal therapy, psychodynamic therapy, non-directive counseling, and various supportive interactions and types of help. Health professionals or the public provides these interventions through individual or group access over the phone, at home, and during clinical visits. Psychosocial interventions are unstructured and not manualized, while psychological interventions are manualized, such as cognitive behavioral therapy, interpersonal therapy, and psychodynamic therapy [20]. Such extensive psychosocial and psychological interventions are expected to support and manage the stressors induced in ICU nurses caring for pediatric patients. It is necessary to consider how it affects them. Therefore, based on the question, “Do ICU nurses caring for pediatric patients who receive psychosocial and psychological interventions show a difference in the levels of stress, distress, depression, anxiety, and trauma than the groups that receive routine care (such as medication by a clinician) or no intervention?” This study performed a systematic review of psychosocial and psychological interventions for ICU nurses caring for pediatric patients.

The significance of evidence-based nursing has been highlighted with a recent growing emphasis on its importance in medicine. Hence, it is crucial to evaluate psychosocial and psychological interventions’ effects in ICU nurses caring for pediatric patients, and the most effective interventions should be identified and administered to them based on these. This will improve their mental health and enhance the quality of nursing care for pediatric patients.

Thus, this study aimed to conduct a systematic review of journal-published articles that have investigated psychosocial and psychological interventions’ effectiveness among ICU nurses caring for pediatric patients to draw broad and objective conclusions on the subject. The specific objectives are as follows: 1) examine psychosocial and psychological interventions’ characteristics provided for ICU nurses caring for pediatric patients and 2) assess the outcome measures and effect sizes of the psychosocial and psychological intervention programs provided for ICU nurses caring for pediatric patients and analyze their statistical significance. This study’s findings illustrate the most effective intervention elements and methods promoting nurses’ psychological adjustment and mental health. They also serve as foundational data for improving pediatric nursing care’s quality.

## 2. Materials and Methods

### 2.1. Study Design

This systematic review analyzed psychosocial and psychological interventions’ effects on ICU nurses caring for pediatric patients. It is a meta-analysis conducted to calculate the effect sizes for the outcome measures.

### 2.2. Inclusion and Exclusion Criteria

The participants, intervention, comparisons, outcomes, the timing of outcome measurement, settings, study design strategy was used in this study. Participants were studies on psychosocial and psychological interventions administered to ICU nurses caring for pediatric patients, and intervention was psychosocial and psychological interventions used. The comparison was routine management (i.e., regular care and standard medical care) or no intervention at all. Outcomes were stress, distress, depression, anxiety, and trauma, and the timing of outcome measurement was the intervention’s duration. Lastly, settings were the intervention’s place of occurrence, and the study design was experimental.

The inclusion and exclusion criteria are given below.

Inclusion criteria: 1) studies on psychosocial and psychological interventions administered to ICU nurses caring for pediatric patients, 2) experimental studies, and 3) studies published in journals.

Exclusion criteria: 1) studies not conducted on ICU nurses caring for pediatric patients, 2) one-group studies without a control group, 3) degree dissertation, 4) reviews, 5) case series, and 6) pilot studies that did not proceed to the next step.

### 2.3. Literature Search and Selection

#### 2.3.1. Search Strategy

This study was conducted according to the preferred reporting items for systematic reviews and meta-analysis (PRISMA). The data were collected from January 1-June 28, 2021. The primary objective when collecting data is to analyze the elements and effectiveness of psychosocial and psychological programs provided for ICU nurses caring for pediatric patients.

Literature searches were performed using PubMed, EMBASE, and CINAHL.

Medical subject headings (MeSH) and EMBASE TREE were used as search terms. The following search terms were combined using “and” as necessary: “Intensive Care Units, Neonatal” (MeSH Terms), “ICU, Neonatal,” “Neonatal ICU,” “Neo-natal Intensive Care Units,” “ Neonatal,” “Newborn ICU,” “Intensive Care Units, Pediatric,” “ICU, Pediatric,” “Pediatric ICU,” “Pediatric Intensive Care Units,” or “Pediatric,” and “Nurse” (MeSH Terms), “Nurse,” “Nurses,” “Psycho-logical Therapy” (MeSH Terms), “Psychological Therapy,” “ psychosocial support system,” “psychological,” “psychology,” “psychosocial,” “psycho-social,” “psychoeducat*,” or “psychoeducat*,” and “Stress Disorders, Traumatic” (MeSH Terms), “depression” (MeSH Terms), “stress,” “distress,” “depression,” “anxiety,” or “trauma.” The searches were limited to human studies, published in the past ten years, and published in English.

#### 2.3.2. Data Selection

Two nursing professors selected and reviewed the data. Each researcher independently reviewed the data and summarized the findings using a uniform format to share during weekly meetings. Ten team meetings were conducted. Each researcher independently performed the literature search and selection. Their selections were reviewed during team meetings. Any disagreement between the researchers was resolved by reviewing the full text until an agreement was reached.

#### 2.3.3. Quality Appraisal

The studies’ quality was assessed regarding their risk level (i.e., high, low, or uncertain) for detection bias, attrition bias, and reporting bias [21]. Two researchers independently assessed each study’s quality and generated a list of the rated studies in codes. A final rating was determined through a discussion between the researchers.

A funnel plot of each study’s estimated effect sizes (horizontal axis) against the sample size (vertical axis) in the RevMan 5.3 software was used to assess the publication bias. In general, ten or more studies were used for the analysis, and the funnel plot’s symmetry was used to assess bias [22].

### 2.4. Data Analysis

#### 2.4.1. Features of intervention programs

Four domains (i.e., study and participant characteristics, intervention, outcome variables, and study outcomes) were analyzed.

#### 2.4.2. Effect size of Intervention Programs

Cochrane’s RevMan version 5.3 software analyzed the intervention programs’ effect sizes and homogeneity. The heterogeneity of the significant variables was tested through null hypothesis testing using the chi-square test. A value of 0% indicates no heterogeneity, while a value of 30%–60% and greater than 75% indicates moderate and considerable heterogeneity, respectively [21].

Forest plots were used to examine the effect’s directions and confidence interval. In contrast, the effect size of the outcome was analyzed using standardized mean differences for continuous data. Statistical significance was set at .05, and a 95% confidence interval was used.

## 3. Results

### 3.1. Data Selection

In the first round of literature search, 1,630 English studies were generated in PubMed (1,040), EMBASE (165), CINAHL (425). Duplicate search results were excluded (111), and the Endnote software was used to manage the searches. In contrast, 672 studies that were not relevant to the study topic and 259 non-experimental studies were excluded in the first round. After reviewing the articles’ full texts, 54 reviews, 51 qualitative studies, and 204 studies in which ICU nurses caring for pediatric patients were not the primary study population were additionally excluded. Hence, four studies were finally included in the analysis (Figure 1).

**Figure 1.**
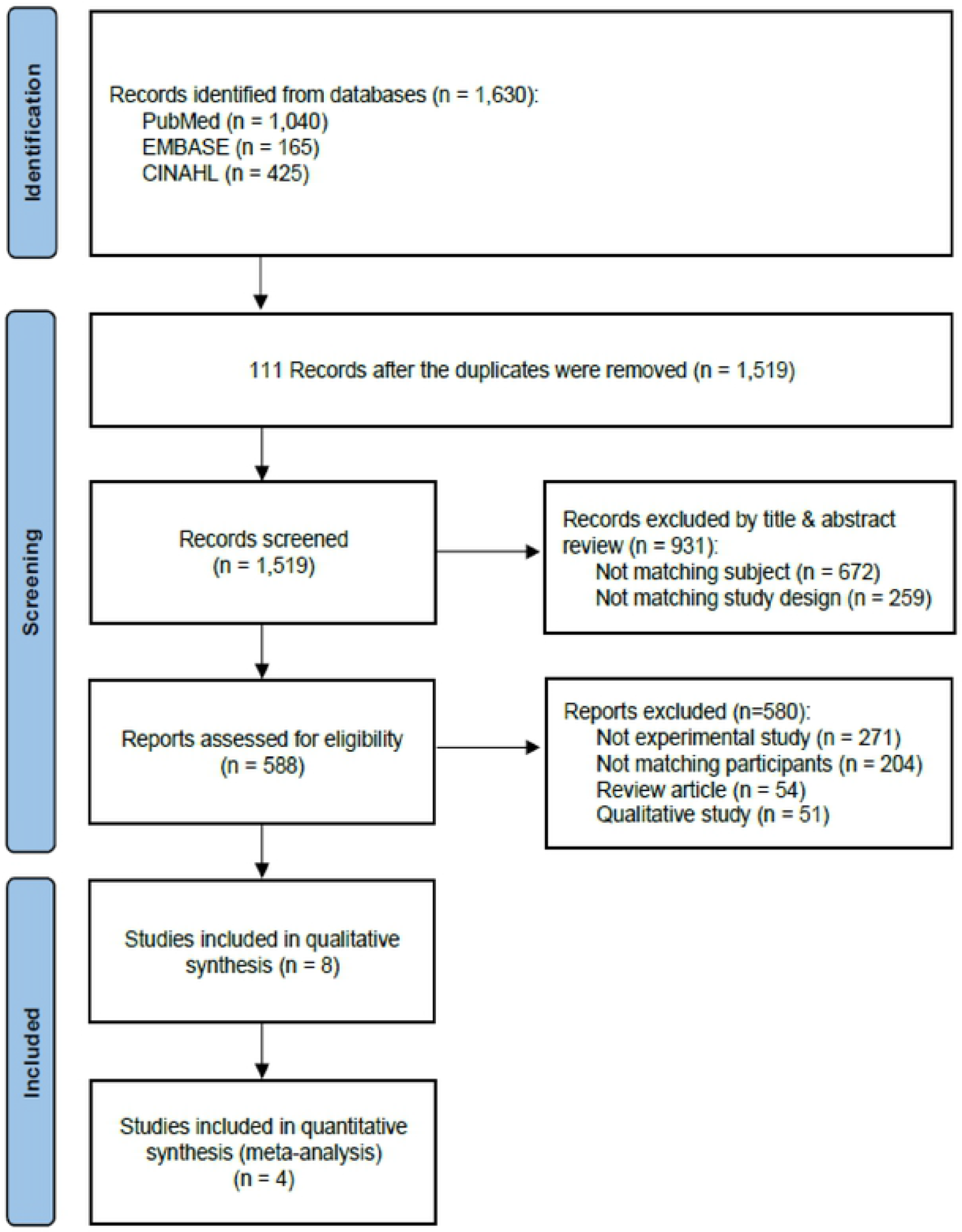
PRISMA flow diagram of the study screening.

### 3.2. Quality Appraisal

To evaluate the quality of the literature included in this study, the risk of bias assessment tool for non-randomized studies were used to assess the quality of non-randomized controlled trials. Bias due to confounding, bias in the subject selection, bias in intervention classification, bias due to deviation from the intended intervention, bias due to missing data, and bias in selecting reported results was investigated [21].

After the quality evaluation for non-randomized controlled trial (n=4) examining the bias due to confounding factors such as whether external events or situational changes at the intervention time that may affect the intervention outcome are experienced equally in both groups, the pre-intervention trends, and whether the patterns of intervention results were accurately analyzed, all four studies were judged to be similar and low in bias. Regarding the bias in the subject selection, there were three studies in which the two groups for comparison were judged to be similar, and the subject assignment was considered appropriate. In one study, compared to nurses and doctors, the selection bias of the comparison group was high. Regarding the bias due to high dropout rates from the intended intervention, the dropout rates of the four studies were 0%, 8.1%, 12.5%, and 31.2%, respectively, indicating a high degree of bias due to dropout rates in some. Regarding the bias in the classification of interventions, examining whether the specialization of the pre-intervention and post-intervention time points influenced the outcome after the intervention revealed no intended intervention bias in all studies. The bias due to missing data was low since all four studies reported the complete collected data. Regarding the bias in selecting reported results, such as if the numerical research results evaluated were selected from various measurements according to the research results, we found that all expected results were reported in all four studies, including those planned in advance.

We did not assess the risk of publication bias in this study because it is recommended to include at least ten studies in assessing publication bias [22].

### 3.3. Characteristics of The Included Studies

The characteristics of the studies that administered psychosocial and psychological interventions to ICU nurses caring for pediatric patients were as follows. Regarding the publication year, two studies were published in 2017 and two in 2019. Regarding the country, there was one Turkish study, two Iranian studies, and one United States study. The study design included a randomized controlled trial (one study), a quasi-experimental study (two studies), and a pretest-posttest intervention study (one study). The study population included neonatal intensive care unit (NICU) nurses (two studies) and pediatric intensive care unit (PICU) nurses (two studies). The pooled sample size was 263 (167 in the intervention group and 141 in the control group). One study used a crossover design with 45 individuals. The psychosocial and psychological intervention types for ICU nurses caring for pediatric patients included aromatherapy (one study), a stress management program (McNamara training; one study), narrative writing (one study), and an Exploration of Patient Ethics and Communication Excellence (PEACE) round (one study). The administration form was individual (one study), group (two studies), or not reported (one study). The outcome measures were stress (two studies), moral distress (two studies), and anxiety (one study). There were no significant differences between the groups in all four studies. However, one study that administered McNamara training, a type of stress management program, led to significant changes in the experimental group. Conversely, another study that administered PEACE rounds reported that the nurse group showed reduced moral distress than the physician group. Hence, two studies reported significant changes in the experimental group after the psychosocial and psychological intervention among ICU nurses caring for pediatric patients (Table 1).

**Table 1.** Descriptive summary of included studies.

| Study | Design /Participants (n) | Intervention | Outcome (Scale) | Results ($p$-value) |
| --- | --- | --- | --- | --- |
| Eren et al. [24]<br>2017<br>Turkey | Quasi experimental clinical study (cross matching groups)<br>ICU (included PICU) nurses (n=45)<br>- Ig: n=45<br>- Cg: n=45 | Type: Aromatherapy<br>Session: Once for 10 to 15 minutes<br>Format: individual approach | Perceived Stress Scale (PSS)<br>State-Trait Anxiety Inventory I -II<br>(STAI- I and II)<br>Visual Analog Scale (VAS/Pleasure)<br>Arterial blood pressure and pulse | Stress: $p = 0.530$<br>STAI- I :<br>$p = 0.348$<br>STAI-II:<br>$p = 0.917$ |
| Lary et al. [5]<br>2019<br>Iran | Quasi-experimental study<br>NICU nurses (n=70)<br>- Ig: n=35<br>- Cg: n=35 | Type: The McNamara education Program (Stress Management program)<br>Session: 1hour sessions for 6 weeks<br>Format: group approach | The Stress Response Inventory (SRI) | Stress response : $p = 0.021$ (Ig),<br>$p =$ not reported (Cg) |
| Saeedi et al. [25]<br>2019<br>Iran | RCT<br>ICU and NICU nurses (n=106)<br>- Ig: 55<br>- Cg: 51 | Type: Narrative writing<br>Session: once a week for 8 weeks<br>Format: group or individual approach not reported | The frequency and intensity of moral distress | Intensity of moral distress:<br>$p = 0.800$<br>Frequency of moral distress:<br>$p = 0.500$ |
| Wocial et al. [26]<br>2017<br>USA | Prepost intervention design<br>Physicians and nurses in PICU (n=42)<br>- RN(Ig): n=32<br>- MD(Cg): n=10 | Type: Pediatric Ethics and Communication Excellence (PEACE) Rounds (formal facilitated discussion)<br>Session: Over a 12-month period, once a week<br>Format: team approach | The Moral Distress Scale Revised (MDS-R)<br>The Pediatric Risk of Mortality (PRISM-3) Score and Risk of Mortality | Moral distress : RN $p = 0.08$ (Ig),<br>MD $p = 0.80$ (Cg) |

Note: ICU = intensive care unit; PICU = pediatric intensive care unit; Ig = Intervention group; Cg = Control group; NICU = neonatal intensive care unit; RCT = randomized control trial; RN = registered nurse; MD = medical doctor

### 3.4. Outcome variables and Effect size

The primary outcome variable in the included studies was stress. Figure 2 presents the results.

**Figure 2.**
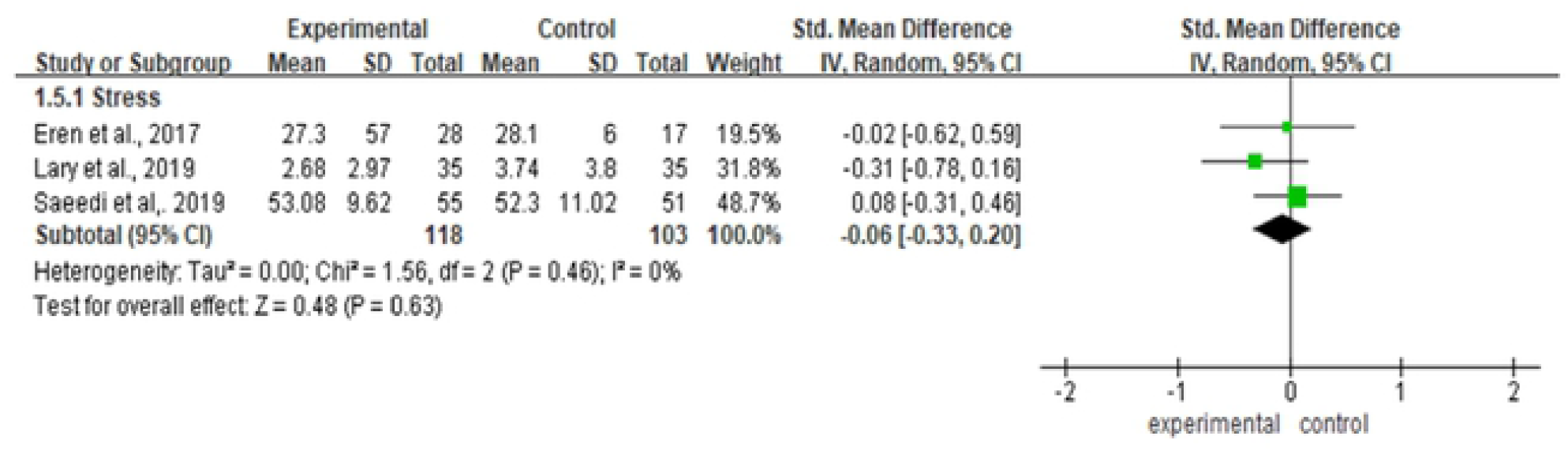
Forest plots of the effect of psychosocial supportive intervention.

#### 3.4.1. Stress

Three studies that reported stress were analyzed. These studies were not heterogeneous, with Q (Chi^2^) = 1.56, df = 2 (p = 0.460); I^2^ = 0%. Moreover, the intervention programs were found to have no effects on stress in each study. The overall effect size was -0.06 (95% CI: -0.33, 0.20), which was not statistically significant (Z = 0.48, p = 0.630).

## 4. Discussion

We performed a systematic review and meta-analysis to examine the psychosocial and psychological intervention methods administered to ICU nurses caring for pediatric patients and integrate these interventions’ effects. The systematic review included a total of four studies published within the past ten years.

The types of psychosocial and psychological interventions for ICU nurses caring for pediatric patients in the four included studies were aromatherapy [24], stress management programs [5], narrative writing [25], and PEACE rounds [26]. Most of these psychosocial and psychological interventions were used as group interventions [5, 26]. Except for one study [25], all studies were non-randomized controlled trials. Psychosocial supportive interventions to manage and prevent negative emotions in ICU nurses caring for pediatric patients should be more actively developed. This is because managing nurses’ negative emotions can directly influence pediatric nursing care’s quality [8, 17] Additionally, most included studies were non-randomized controlled trials. Thus, additional randomized controlled trials should be conducted to present study findings with a higher evidence level.

The outcome measures examined by the four included studies were stress [5, 24], moral distress [25, 26], and anxiety [24]. This shows that most studies examined stress and moral distress. Further, NICU and PICU nurses experience negative emotions such as psychological stress, moral distress, depression, and guilt from the burden of providing nursing care for weak children, pediatric patients’ death, inappropriate coping skills, and ethical dilemmas encountered during pediatric nursing care [8, 10, 13]. These influences nurses’ affection and compassion, thereby deteriorating pediatric nursing care’s quality [8, 17]. Thus, active support should be provided for ICU nurses caring for pediatrics so that situations that may trigger negative emotions or consequent negative emotions can be converted into more positive emotions.

Regarding the psychosocial and psychological interventions’ effectiveness for ICU nurses caring for pediatrics, there were no significant differences between the two groups in any of the four studies [5, 24-26]. However, one study that administered a stress management program to NICU nurses [5] and one that administered PEACE rounds to PICU nurses and physicians [26] reported significant changes within the experimental group. These results align with a previous pilot study’s finding that administered a mindfulness-based intervention to 38 PICU nurses with no significant differences in stress between the two groups. However, there were significant changes in stress levels within the groups over time [27].

In the four included studies, one study administered the McNamara education program (a stress management program) on NICU nurses. This study observed significant changes in the experimental group’s stress response after the program [5]. In contrast, one study that administered the PEACE rounds, which involved structured discussions, on PICU nurses and physicians observed significant changes in the nurse group’s moral distress [26]. This suggests that it is vital to examine appropriate outcome measures for the intervention type applied to the participants. Specifically, there are three steps to confirm that the interventions are effective. First is identifying the negative emotion that requires the most critical intervention among the target population. The second is providing the intervention that helps alleviate such an emotion. The last step is examining relevant outcome measures.

The interventions did not affect the stress in the meta-analysis of the three studies. However, a previous study that administered a stress management program to NICU nurses reported that the experimental group showed significant changes in their stress response [5]. In this study, one out of three studies used a group approach [5], and one used an individual approach [24]. The third study did not mention the approach type [25]. However, it may have used an individual approach because the intervention was narrative writing. Furthermore, although not included in the meta-analysis, one study included in the systematic review also used a team approach (PEACE Rounds) [26]. In this study, there were significant changes in the nurse group’s moral distress after the intervention. A previous study that administered a group-based mindfulness intervention for PICU and NICU healthcare providers reported significant changes in depression and posttraumatic stress disorder symptoms within the group [28]. Another study that applied a group-based mindfulness therapy to PICU nurses reported significant changes in the group’s stress levels over time [27]. In contrast, an individual approach using mindfulness therapy for novice pediatric nurses did not lead to significant changes in stress and posttraumatic stress disorder [29]. Moreover, a narrative writing intervention administered to ICU nurses, including NICU nurses, did not significantly change moral distress’ intensity and frequency [25]. Additionally, an individual approach using aromatherapy for ICU nurses, including PICU nurses, did not lead to significant changes in stress and anxiety [24]. Thus, these results suggest that using an individual or group approach significantly impacts the interventions’ effectiveness. Hence, subsequent studies should primarily implement group approaches and re-analyze their effectiveness since a group approach is speculated to be more effective than an individual approach.

These results show that psychosocial and psychological interventions for ICU nurses caring for pediatrics should be developed after identifying their primary psychosocial and psychological needs. Group approaches are expected to be more effective than individual approaches. Furthermore, as opposed to pretest-posttest experimental studies, long-term follow-up studies should be conducted to examine the varying effects on outcome measures over time.

This study had several limitations. The study findings should be interpreted with caution because only studies published in the past ten years in English were included. In the present study, only four studies met the inclusion criteria. Despite these limitations, this study is significant because it is the first to assess the psychosocial and psychological interventions’ effectiveness for ICU nurses caring for pediatrics. Moreover, it presented implications for future studies by examining intervention types and effectiveness based on the latest evidence available.

## 5. Conclusions

This study aimed to analyze psychosocial and psychological interventions’ elements and effects provided for ICU nurses caring for pediatric patients. Four studies that met the inclusion criteria were selected after reviewing 1,630 studies. The interventions used in these studies primarily targeted stress management, including moral distress. However, they were ineffective in reducing stress and moral distress. Stress, including moral distress, is an emotional domain that may be influenced by individual sensitivity and the work environment. People may struggle with altering their negative emotions in a short span. Thus, supportive interventions should be developed based on an analysis of the pediatric critical care unit’s nurses’ psychosocial and psychological states and work environments. To this end, the standard negative emotions triggered during pediatric care should first be examined. These can be used to develop various psychosocial supportive intervention programs that help detect and rationally process such emotions or those that help positively perceive such negative emotions and aid in positive coping strategies.

The effect size in this meta-analysis should be interpreted with caution because only four studies were included, and most of them were non-randomized controlled trials. Therefore, subsequent studies should assess the effectiveness of high-quality interventions based on well-designed randomized controlled trials with adequate sample sizes.

## Author Contributions

Conceptualization, M.-H.C. and M.L.; Data curation, M.-H.C. and M.L.; Formal analysis, M.-H.C. and M.L.; Investigation, M.-H.C. and M.L.; Methodology, M.-H.C. and M.L.; Project administration, M.-H.C. and M.L.; Validation, M.-H.C. and M.L.; Visualization, M.-H.C. and M.L.; Writing—original draft preparation, M.-H.C. and M.L.; Writing—review and editing, M.-H.C. and M.L. All authors have read and agreed to the published version of the manuscript.

## Acknowledgements

We would like to thank Editage (www.editage.co.kr) for English language editing.

## Institutional Review Board Statement

Not applicable.

## Informed Consent Statement

Not applicable.

## Data Availability Statement

The data presented in this study is available upon request of the respective author.

